# An AI-Based Chatbot to Support Health-Related Social Needs among Pediatric Primary Care Population: Protocol for a Pilot Randomized Controlled Trial

**DOI:** 10.1101/2025.11.17.25340433

**Authors:** Emre Sezgin, Elizabeth Clarkson, Faith Logan, Daniel I. Jackson, Syed-Amad Hussain, Joshua Stokes, Alicia Bunger, Guy Brock, Eric Fosler-Lussier, Alex R. Kemper, Ahna L. Pai

## Abstract

Unmet health-related social needs (HRSNs) are major drivers of poor health outcomes in early childhood. Children with unmet HRSNs are at greater risk for developmental delays, caregiver stress, and increased healthcare utilization, yet current screening approaches in pediatric primary care are resource-intensive and inconsistently implemented. AI-powered chatbots (conversational agents or virtual assistants) may offer a private, secure, scalable, and cost-effective alternative for identifying unmet needs and connecting families to services. This protocol describes a pilot randomized controlled trial designed to evaluate the feasibility, acceptability, and usability of DAPHNE, an AI-driven chatbot developed to facilitate the identification of unmet HRSNs and provide personalized community resource referrals. One hundred caregivers of children under two years of age will be recruited from Nationwide Children’s Hospital pediatric primary care clinics and randomized to either the standard care (control) group or DAPHNE+ Standard care (intervention) group (n=50 each arm). Caregivers will complete surveys at baseline, 1 month, 3 months, and 6 months post-intervention (depending on the measure). For the intervention group, participants will receive weekly chatbot prompts and on-demand access throughout the 6-month study period. Primary outcomes include study feasibility (recruitment, retention, and survey completion across both arms), acceptability (caregiver-reported ratings in both arms and intervention-specific ratings), and usability of the DAPHNE chatbot (System Usability Scale among intervention participants).

Secondary outcomes include caregiver-reported outcome measures (caregiver stress, self-efficacy, satisfaction with resource access, quality of life), and electronic health record-derived measures (including documentation of HRSN screening and referrals, adherence to well-child visits, missed appointments, emergency department utilization, and estimated healthcare costs). In addition, ten primary care providers will also participate to assess workflow integration and report on current HRSN practices. Mixed-methods analyses will integrate survey data, chatbot engagement metrics, and qualitative interviews to refine both the intervention and the study protocol. The results of this study will inform the design of a future multi-site trial to evaluate the efficacy and implementation of DAPHNE for addressing HRSNs in pediatric primary care. Trial registration: NCT07168382.

## INTRODUCTION

Health-related social needs (HRSNs), including food insecurity, unstable housing, transportation challenges, and financial instability, account for more than 50% of modifiable health outcomes.[1,2] Children with unmet HRSNs have substantially elevated risks for cognitive, behavioral, and developmental difficulties, having greater risk than peers from resource secure households.[3–6] They are also 18% more likely to have emergency department (ED) visits and 36% more likely to experience hospitalizations compared with children without social risks (IRR=1.18, 95% CI [1.12- 1.23]; 1.36, 95% CI [1.26–1.47]).[7] This burden is especially pronounced among infants younger than 2 years, for whom early-life HRSNs, such as food insecurity or benefit instability, are linked to measurable behavioral and regulatory difficulties by six months of age (aOR= 1.64-1.86 across domains; aOR= 2.16 for 2 or more identified HRSNs).[8] This early-life concentration represents a high-yield prevention target and reinforces primary care as the locus for proactive screening and linkage to services.[7–9]

Pediatric primary care clinics (PCCs) are therefore critical access points for early identification and intervention but current screening practices are constrained by staffing shortages, workflow inefficiencies, and high operational costs.[10] Caregivers are also often reluctant to disclose needs due to stigma, language barriers, or confusion about service navigation, which further exacerbates under-identification.[11] Referral completion is constrained by a combination of structural, provider-level, and caregiver-level barriers. Structural barriers include complex eligibility requirements, poor coordination, and limited availability of community resources.

Provider-level barriers, such as limited familiarity with available services and uncertainty about referral processes, further hinder follow-through. Caregivers also face barriers related to time, transportation, and competing demands that make service access difficult.[12–14]

Despite the availability of standardized screening instruments, fewer than 25% of hospitals and 16% of physician practices routinely screen for HRSNs.[15] When implemented, screening is often limited to episodic encounters such as annual well-child visits, missing acute and urgent social needs at other timepoints.[16,17] In this cohort, particularly the caregivers of infants (ages 0–2) are disproportionately affected by the lack of screening.[3,8] For instance,food insecurity in this age group is strongly associated with adverse developmental outcomes,[18] higher hospitalization rates,[19] and increased risk of chronic conditions including obesity and diabetes.[20] The barriers are particularly pronounced for Medicaid-insured children, who experience greater challenges in securing basic necessities such as infant formula, diapers, and transportation.[21,22]

### Rationale for AI-Driven Solutions

AI-based chatbots (also known as conversational agents and virtual assistants) offer an opportunity to overcome some of these structural and provider-related barriers in HRSN identification and referral. Unlike traditional clinician-initiated screening, chatbots can provide continuous, user-initiated engagement across multiple contexts and time points, enabling real-time identification of urgent needs.[23–25] By facilitating private conversations, chatbots play a positive role to reduce stigma and support honest disclosure for some groups.[11,26–28] They can also automate referrals using up-to-date community resource databases, and reduce provider burden while offering tailored, context-specific recommendations.[25,29,30] These advantages are further supported by our preliminary engagement sessions with social work teams, caregivers, and patient advocates, who indicated high acceptance of chatbot-based social needs assessment, provided that privacy and usability are prioritized.[23,31]

### Conceptual Framework

The methods for this study were informed by the Obesity-Related Behavioral Intervention Trials (ORBIT) model[32,33], which provides a systematic framework for developing behavioral interventions by progressing through iterative phases (from early conceptualization and definition of intervention components, to feasibility and pilot testing, and ultimately to efficacy and effectiveness trials). Prior applications of conversational AI in healthcare, including tools for mental health support, symptom tracking, and caregiver-facing applications, demonstrate early evidence and implications for digital engagement strategies.[34–39] Building on this foundation, the present pilot randomized controlled trial (pRCT) corresponds to Phase IIb of ORBIT, which emphasizes feasibility testing, establishing acceptability, and refinement of study protocols before progression to full-scale efficacy trials.

### Study Objectives

The long-term goal of this research is to develop an empirically supported, scalable, AI-driven solution to facilitate HRSN identification and referral in pediatric primary care. This pilot study has three specific objectives: (1) To assess the feasibility, acceptability, and usability of the DAPHNE chatbot with caregivers and providers of pediatric primary care patients compared with standard care; (2) To evaluate the feasibility and acceptability of the study protocol, including recruitment, retention, and measurement strategies, for informing design of a future large-scale trial; and (3) To characterize the current standard of care for addressing unmet HRSNs, thereby informing selection of appropriate comparator conditions and outcomes in future trials.

### Preliminary Work

We have conducted several studies to inform DAPHNE’s development. A scoping review of patient-facing chatbots and voice agents revealed high interest but a lack of rigorous trials on chatbot-based interventions and indicating the need for systematic evaluation.[40] Building upon this knowledge, we developed a voice-interactive diary for caregivers of children with special health care needs; over 80% reported ease of use and approximately half increased health tracking frequency, demonstrating caregiver receptivity to conversational tools.[39] To enable integration of unstructured caregiver data, we created a natural language processing pipeline that accurately structured caregiver-generated notes and transcripts (F1 >0.7), validating real-time data capture for clinical use.[41,42] Finally, in a feasibility study of a semi-functional DAPHNE prototype (n=13), community health and social workers rated usability at 72/100 on the System Usability Scale and emphasized its value for triaging referrals.[23] Together, these studies establish feasibility, provider and caregiver acceptance, and the need for a pilot randomized controlled trial.

## METHODS

### Study Design and Setting

This study is designed as a pilot randomized controlled trial (pRCT) guided by the ORBIT model (Phase IIb).[32,33] One hundred caregivers of children ≤2 years of age will be recruited from Nationwide Children’s Hospital Primary Care Network (NCH PCN). Following baseline assessment, participants will be randomized in a 1:1 ratio to either the standard care group or DAPHNE + standard of care group (n=50 per arm). In addition, 10 providers will be recruited to complete surveys on workflow integration and HRSN practices.

The trial will be conducted in NCH PCCs, which provided nearly 250,000 visits in 2024 for mostly Medicaid-enrolled patients. The PCN includes 14 clinics across the Columbus metropolitan area. For the purpose of this study, we identified 2 clinics with the capacity to support the research project and a relatively higher number of patients with unmet HRSNs.

### Study Status and Timeline

This study is currently in the Initiation and Planning phase, with the protocol finalized and Institutional Review Board (IRB) approval obtained (NCH IRB #00004369). We initiated the process of participant recruitment in Q1, Year 1, and are actively enrolling participants. The time schedule is provided in Figure 1. We project that participant recruitment will be completed in Q2 of Year 2, primary data collection (6-month follow-up) will be completed in Q1 of Year 3, and final results are expected for manuscript submission in Q4 of Year 3.

**Figure 1.**
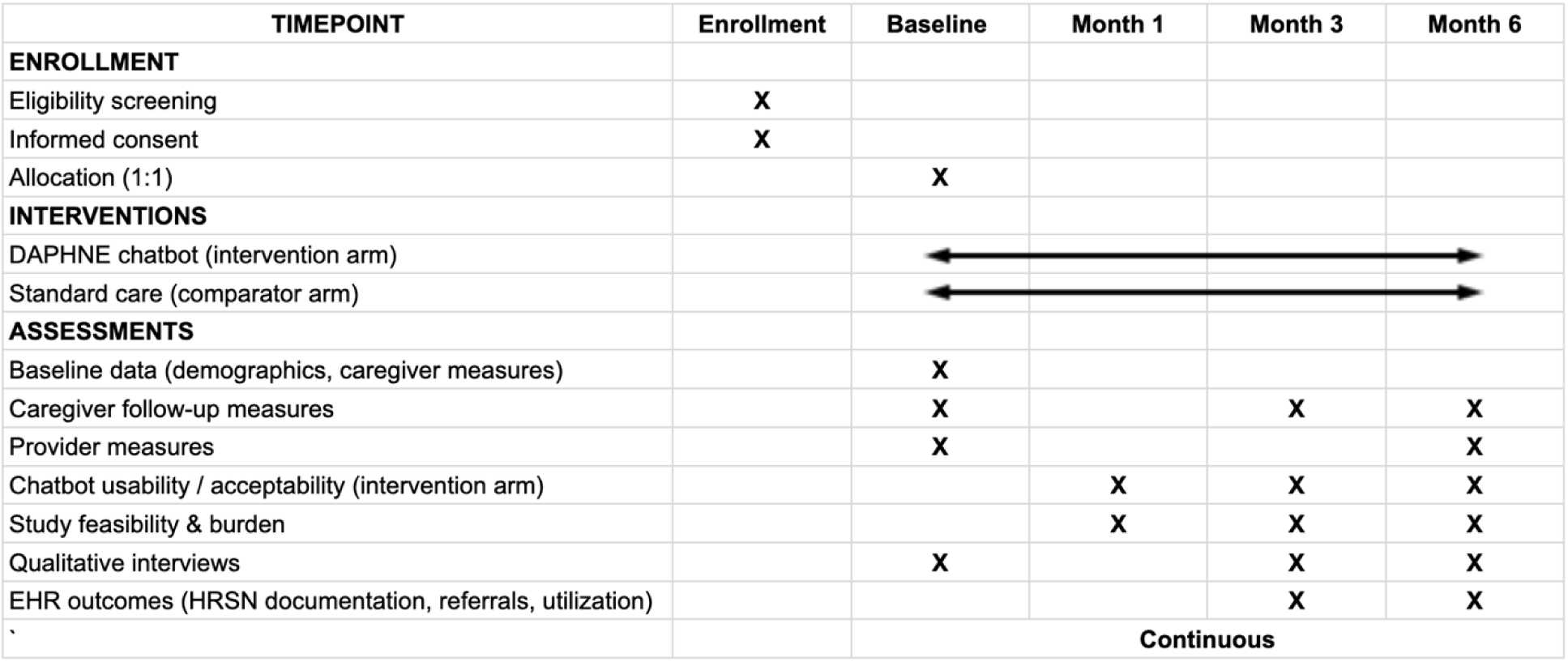
Time schedule of enrolment, interventions, and assessments on participant outcome (informed by the SPIRIT 2025 reporting guideline)[43]

### Participants

*Caregivers.* Recruitment will occur through the NCH PCN, with research staff approaching caregivers during routine visits. We estimate to recruit n=50 participants per arm. We facilitate recruitment using flyers and social worker referrals. Furthermore, we will communicate with potential participants who are interested via virtual methods (phone call, teleconference).

Inclusion criteria are: (1) primary caregiver of a child ≤2 years old receiving care at the selected clinics in the PCN, (2)self-identify with at least one unmet HRSN (3) ability to provide informed consent and complete study surveys in English, (4) ownership of a mobile device for chatbot use, and (5) willingness to participate in follow-up assessments. Exclusion criteria: none beyond failure to meet the above criteria. Caregivers will be compensated for their participation.

*Providers.* To evaluate the feasibility of integrating the DAPHNE chatbot into clinical workflows at primary care clinics, we will recruit approximately 10 providers across the NCH PCN, ensuring representation from at least 50% of the 14 PCC sites. Eligible providers include physicians, nurses, social workers, and care coordinators who are directly involved in patient care. Providers on extended leave or those not engaged in patient care will be excluded.

Recruitment will be facilitated through the NCH Primary Care Research Network.

### DAPHNE Chatbot Development and structure

DAPHNE was developed using secure cloud infrastructure (via AWS). At the backend, we integrated state-of-the-art large language models (e.g. Claude, GPT, Mistral, Gemini, LLaMA) adapted for healthcare settings and enabling intent recognition, contextual follow-up, and dynamic retrieval of resources from the resources API. Unlike rule-based decision trees, the AI allows free-text input from caregivers, interpreting varied language patterns (e.g., “I can’t get to the clinic” means transportation needs) and providing tailored responses. It demonstrated >99% intent accuracy over 1,500 turns of conversations.[31]

DAPHNE is provided as a web-based conversational agent accessible on iOS, Android, or web browsers (Figure 2). It is deployed using HIPAA-compliant servers and validated for accuracy and security by the internal Information Services governance committee. The chatbot will not require computational power on the user’s side, therefore a simple smartphone could be used to engage with the app. Our conversational model is also optimized for scaling at the cloud side with minimal cost and computational resource needs.

**Figure 2.**
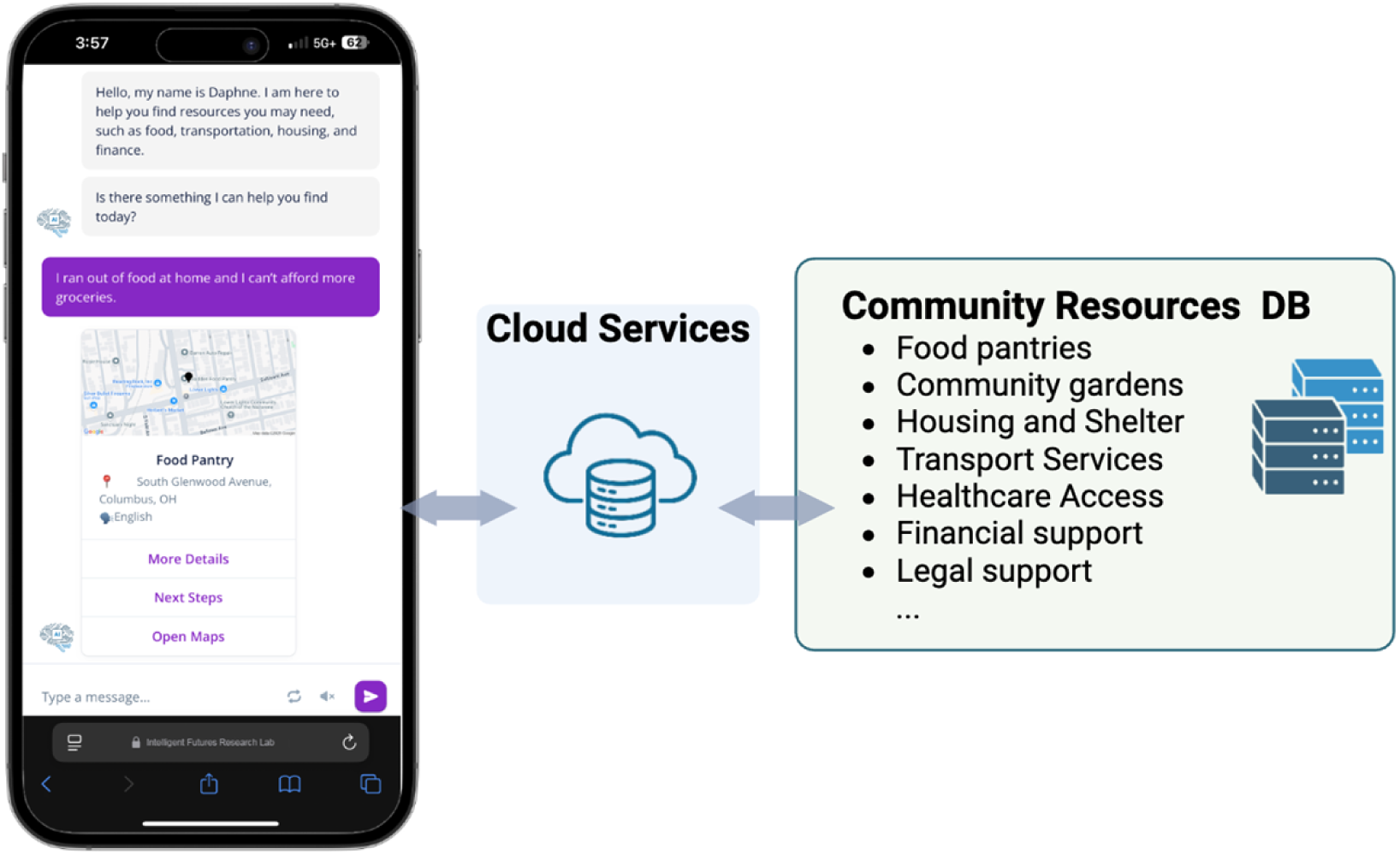
Overview of how the DAPHNE chatbot connects users to community resources

To ensure safety, DAPHNE automatically flags out-of-scope or potentially concerning interactions, and all conversations are subject to monitoring and auditing protocols. The research team reviews flagged transcripts to prevent harmful content and safeguard patient well-being. In addition, the research team periodically reviews the chatbot logs. For urgent or high-risk needs, an escalation protocol is used directing caregivers to professional resources, while the research team coordinates with the social work team as appropriate.

### Intervention: DAPHNE Chatbot

Caregivers randomized to the intervention arm will receive onboarding consisting of instructions on how to download the app and a brief tutorial on the app’s features. The chatbot engages caregivers in natural conversations to understand HRSNs, provide tailored resource recommendations (e.g., food pantries, housing, transportation), and follow-up with automated reminders. Features include periodic personalized notifications (e.g., informational quick tips, updates), information about local services, and guidelines to sustain engagement.

### Control: Standard Care

Caregivers in the control arm will receive the existing clinic-based HRSN screening and referral procedures as currently implemented, without access to the chatbot. These caregivers will complete the same study assessments as the intervention group except those related to chatbot usability and feasibility of the intervention.

### Procedures

All caregivers will complete caregiver-reported outcome measures (cPROMs; see “secondary outcomes” section below) at baseline, 3 months, and 6 months post-intervention. Demographic information will be collected from all caregivers at baseline. Intervention group participants will have on-demand access to the chatbot throughout the 6-month study period, and will receive weekly notifications from the chatbot app. 10 caregivers from both groups will also complete a qualitative interview at 3 months and 6 months to evaluate their experience using the chatbot app versus receiving the standard of care.

### Provider Component

After receiving a brief demonstration of the DAPHNE chatbot, providers will share their feedback and complete a structured survey on current HRSN screening and referral practices. Providers will complete the Workflow Integration Expectancy (WIE) tool and the Feasibility of Intervention Measure (FIM) at baseline and 6 months; providers will also complete the SUS at baseline.

Additionally, providers will complete a survey on the feasibility of implementing the chatbot into their clinical workflows at baseline and 6 months. Semi-structured interviews will be conducted to elicit feedback on integration challenges, potential facilitators, and perceived impact on team workflow. Data from provider surveys will be analyzed descriptively, and interview transcripts will undergo thematic analysis to identify patterns in feasibility, barriers, and opportunities for scaling.

### Outcomes and Measures

*Primary outcomes (Quantitative)*

The primary outcomes will evaluate study feasibility (recruitment, retention, and survey completion across both arms) and intervention usability/acceptability. Feasibility will be assessed through enrollment rates (target ≥ 70%), retention rates (target ≥ 70%), and implementation feasibility using the Feasibility of Intervention Measure (FIM; target ≥ 80%).[44,45] Acceptability will be measured with the Website Evaluation Questionnaire (WEQ), with success defined as ≥ 80% endorsement of top rating categories.[46] Usability will be assessed using the System Usability Scale (SUS), with a mean score ≥ 68 considered acceptable.[47] Technical performance will also be evaluated, including chatbot response accuracy (target F1 ≥ 0.7 [48]), response latency (<3 seconds), and differences across platforms (iOS vs. Android).In addition, caregiver comprehension will be assessed using the Patient Comprehension Questionnaire (PCQ), with success defined as a mean score ≥4.The PCQ will be administered to caregivers in the intervention arm at baseline and 3 months.

*Primary outcomes (Qualitative)*

To further assess the DAPHNE chatbot, qualitative analysis will be completed for caregiver and provider interviews. This qualitative data will overlap with quantitative measures to gain a stronger understanding of provider and caregiver perspectives on feasibility, acceptability, and usability of the DAPHNE chatbot. For caregivers in the intervention group, qualitative interviews will be coded to evaluate factors that drive caregiver engagement with the DAPHNE chatbot and encourage use of the chatbot as well as barriers that may impact caregiver ability or want to use the chatbot. Interviews for caregivers in the control group will provide information about the current standard of care for caregivers with social needs and allow a comparison between resources or suggested resources caregivers receive in the current standard of care versus resources or suggested resources caregivers received while using the chatbot. Caregiver interviews across both groups will also be evaluated for caregiver perspectives toward study burden to determine the acceptability and feasibility of completing a larger scale trial of the DAPHNE chatbot. Clinician interviews will give first-hand insight into how well the DAPHNE chatbot could be integrated into clinic workflow by focusing on how useful the chatbot is perceived to be in a clinic setting, the challenges that may arise from using the chatbot in a clinic setting, and the ease with which the chatbot would be able to be integrated and possibly enhance the current standard of care.

*Secondary outcomes*

Secondary outcomes address caregiver- and provider-level effects. Caregiver-reported outcomes (cPROMs) include stress,[49] self-efficacy,[50] satisfaction with access to community resources,[51] and quality of life for caregiver and child as proxy.[52] These cPROMs will be completed by caregivers in both arms at baseline, 3, and 6 months. In addition to cPROMs, caregivers in both arms will complete the standardized HRSN screener. This survey will be administered at 3 and 6 months to capture changes in reported needs over time. Caregiver engagement with the chatbot is measured through system logs (e.g., minutes of use, number of logins, session durations, screenings completed, and resources accessed).

Caregiver burden associated with study procedures will be measured in both arms using the Participant Burden Assessment (PeRBA; target ≥4) at 1, 3, and 6 months.[53] Provider perspectives on workflow integration will be assessed using the Workflow Integration Expectancy tool, with a target rating ≥ 4.[54] These secondary outcomes will be analyzed to see how they change over the course of the trial and to eventually address whether there are any associations with outcome changes and chatbot use.

*Exploratory outcomes*

Exploratory analyses will focus on healthcare utilization and economic outcomes to inform future large-scale trials. Electronic health record (EHR) data will be extracted to capture documentation of HRSN screening and referrals, adherence to well-child visits, missed appointments, emergency department utilization, and estimated healthcare costs. EHR data will be abstracted for all participants at 3- and 6-month follow-up period to capture pediatric primary care visits, social work referrals, and emergency department utilization. Billing and visit data will be used to estimate healthcare costs, including cost avoidance from reduced no-shows or ED visits. While the study is not powered to detect cost effects, descriptive comparisons between the DAPHNE and standard care arms will be conducted to identify patterns in service utilization and associated costs. Findings will inform the selection of economic outcomes and power calculations for a future R01 trial. Additional exploratory outcomes will include continuous tracking of recruitment rates and dropout rates throughout the study period. Outcome measures, descriptions and planned timeline are also available at our registered protocol.[55]

### Sample Size Justification

The target enrollment of 100 caregivers (50 per arm) is appropriate for a pilot trial. The study is not powered for efficacy testing; rather, the focus is feasibility, acceptability, and usability outcomes to inform the design of a future fully powered R01 trial. The sample size provides a precision (half-width of 95% confidence interval) of 0.4 standard deviations for estimating mean differences between groups.

### Data Analysis

Quantitative data (survey scores, engagement metrics, retention rates) will be summarized using descriptive statistics (e.g., mean, median, SD and IQR). Qualitative data will be transcribed and analyzed using thematic content analysis (hybrid inductive-deductive approach) in NVivo by two independent coders with ≥85% inter-rater reliability. Integration of quantitative and qualitative findings will follow a convergent mixed-methods design,[56,57] in which both data streams are analyzed separately and then merged during interpretation. Integration will occur through joint display and narrative weaving, identifying points of convergence, divergence, and complementarity to refine the intervention and inform future trials. Exploratory analyses will compare intervention and control arms on secondary outcomes (caregiver-reported stress, self-efficacy, satisfaction; EHR-derived service use) using t-tests or Wilcoxon rank-sum tests and descriptive comparisons. Effect sizes will be estimated to inform future power calculations.

### Randomization and Analysis Population

Participants will be randomized 1:1 to DAPHNE or standard care using a pre-generated allocation sequence from the NCI Clinical Trial Randomization Tool (maximal procedure; maximum tolerated imbalance = 2). Allocation concealment will be maintained by an unblinded coordinator who releases the assignments via secure research tool (RedCAP) to be shared with the study team during recruitment. The primary analytic approach will follow the intention-to-treat (ITT) principle, whereby all randomized participants are analyzed in their originally assigned group regardless of adherence or withdrawal. This ensures comparability between arms and reflects real-world implementation.

### Missing Data

For feasibility endpoints (e.g., retention, completion), denominators will include all randomized participants; nonresponders will be classified as not completing that timepoint. For continuous outcomes (e.g., SUS, WEQ, PCQ), available-case summaries will serve as the primary analysis, with prorating applied when allowed by instrument scoring manuals. If missingness exceeds ∼10–20% for a given outcome, multiple imputation using chained equations will be conducted as a sensitivity analysis, incorporating arm, site, baseline measures, and predictors of missingness. Engagement metrics for the intervention arm will be analyzed under ITT, with non-users coded as zero activity to avoid survivorship bias. Sensitivity analyses will compare complete-case, imputed, and worst-case assumptions to evaluate robustness.

### Data Monitoring and Risk Management

All study data will be collected through HIPAA-compliant electronic systems and stored on encrypted Nationwide Children’s Hospital servers. Data integrity will be ensured through automated validation checks, routine audits, and adherence to standardized abstraction protocols. Technical support will be available via a dedicated helpdesk during working hours, with all issues logged for iterative improvement.

Potential risks include participant frustration due to usability issues, privacy concerns, or inaccurate chatbot responses. To address these risks, the study incorporates caregiver training during onboarding, visual and video guides, and reassurance that DAPHNE complements rather than replaces clinical care. The chatbot’s performance will be continuously monitored for response accuracy and timeliness, with misinformation or errors flagged and corrected during weekly team reviews. Provider oversight will be integrated through periodic reporting of chatbot-identified needs and referrals to ensure appropriate follow-up. These strategies are designed to maintain caregiver trust, ensure accurate outputs, and safeguard participants throughout the study.

An independent Data Safety and Monitoring Board, consisting of 4 experts in primary care practices, AI/ML research, and biostatistics will oversee the trial to ensure participant safety, data integrity, and overall trial conduct. The board, which is independent of the study sponsor and funders, will meet semi-annually to review cumulative safety data, enrollment metrics, and protocol adherence. They will provide recommendations directly to the Principal Investigator and the managing institution regarding the continuation, modification, or termination of the study.

## Discussion

This protocol describes a pilot randomized controlled trial of DAPHNE, an AI-driven conversational agent designed to improve identification of health-related social needs and facilitate caregiver access to essential community resources. The study has potential to address the access barriers in pediatric primary care, where unmet HRSNs remain a major driver of adverse child health outcomes, yet routine screening and referral processes are underutilized.[15–17]

By focusing on feasibility, acceptability, and usability, this study will generate critical data to guide refinement of both the intervention and the study protocol. Specifically, it will assess caregiver and provider engagement, evaluate technical performance across devices, and characterize barriers to study participation. Complementary analyses of provider surveys and EHR data will contextualize the current standard of care, providing essential information for selecting comparator conditions and outcomes in future clinical trials.

The results will directly inform a larger, multi-site R01 trial evaluating the efficacy and implementation of AI-driven HRSN screening. Consistent with the ORBIT model, [32,33] this study represents an intermediate phase (IIb) in behavioral intervention development, building the foundation for robust testing of clinical and operational outcomes. Longer-term directions include expansion to non-English-speaking populations, integration of voice interactivity, and cost-effectiveness analyses to assess healthcare savings from reduced no-shows and emergency utilization.

If successful, DAPHNE could advance personalized, equitable pediatric care by enabling real-time HRSN screening, reducing stigma in caregiver disclosures, and streamlining referral pathways. Beyond pediatrics, this approach has the potential to provide a generalizable framework for integrating AI-driven conversational tools into routine healthcare delivery and public health systems.

### Strengths and Limitations

This study has several important strengths. First, DAPHNE introduces an innovative, AI-driven approach to health-related social need (HRSN) screening, using natural language, private, and context-aware interactions that move beyond static questionnaires.[23–25,29,30] Second, the intervention was co-designed with caregivers, clinicians, and advocates, ensuring that usability, engagement, and contextual relevance are embedded in its design.[23] Third, the study emphasizes integration into pediatric primary care workflows, which is essential to reduce burden on care teams while maintaining alignment with provider responsibilities.[58,59] Fourth, the trial is guided by the ORBIT model,[32,33] which provides a systematic framework for developing and iteratively evaluating the DAPHNE behavioral intervention. Finally, the modular design of backend resource integration allows integration with diverse community resource databases, making the intervention scalable and adaptable to local or health-domain specific populations and clinical contexts.[60–65]

Nonetheless, there are key limitations. This is a single-site pilot study with a relatively small sample size (n=100) from specific region/ state, which restricts the generalizability of findings and precludes formal efficacy testing. Results will instead focus on feasibility, acceptability, and usability outcomes to inform future large-scale trials. The study is also limited to English-speaking, smartphone and data plan owner caregivers, which constrains equity in early testing. The 6-month follow-up period may not capture longer-term outcomes such as sustained engagement, resource utilization, or downstream effects on clinical care and health outcomes. Finally, technology-specific challenges, including variable device access, inconsistent internet connectivity, and risks inherent to conversational AI (such as inaccurate responses or hallucinations), may influence feasibility and participant trust. [66–69]

Even though we hypothesize the benefits of chatbot, such as the automation of referrals, increased utilization of resources and the potential reduction of provider burden by efficiently identifying HRSNs, we invite readers and practitioners to be cautious. There are implicit factors including potential AI biases, AI related errors and inaccuracies and human overreliance on AI for resource support, which may potentially limit implementation of chatbot at a scale.

Technology in our project focuses on supplementing human support rather than replacing, therefore not diminishing the value of personalized assistance and human interaction in practice.[70]

## Conclusions

This pilot randomized controlled trial will provide essential feasibility, acceptability, and usability data on the DAPHNE chatbot for addressing health-related social needs in pediatric primary care. Findings will inform refinement of both the intervention and the study protocol, directly supporting the design of a future multi-site R01 trial. Guided by the ORBIT framework, the study emphasizes early-phase testing as a foundation for scalable, rigorous evaluation. In future research, the trial will expand to larger and more diverse populations, incorporate non-English language options and voice interactivity, and integrate cost-effectiveness analyses to assess the impact on healthcare utilization. Ultimately, this work will determine whether AI-driven conversational agents can serve as a sustainable and equitable model for integrating social needs identification and referral into routine pediatric care.

## Data Availability

No datasets were generated or analysed during the current study. All relevant data from this study will be made available upon study completion.

## Acknowledgements

We gratefully acknowledge the support from the AWS Health Equity Initiative (via AWS Social Responsibility and Impact credits), as well as the FindHelp.org Fellowship to maintain community resource navigation. Our appreciation is also extended to the NCH IT-RI team for their expertise in platform development and to the NCH Office of Technology Commercialization for their guidance in scaling and dissemination.

## Ethics

The study has Institutional Review Board (IRB) approval at Nationwide Children’s Hospital (#00004369). Informed consent will be obtained from all participants.

## Data Availability Statement

Not available

